# Identifying drug repositioning candidates for age-related outcomes in the Danish health registries

**DOI:** 10.1101/2024.08.12.24311869

**Authors:** Alexander Wolfgang Jung, Ioannis Louloudis, Søren Brunak, Laust Hvas Mortensen

**Affiliations:** Department of Public Health, Faculty of Health and Medical Sciences, University of Copenhagen, Copenhagen, Denmark; Statistics Denmark, Copenhagen, Denmark

**Keywords:** Real-World Evidence, Drug repositioning, Targeted minimum loss estimation (TMLE)

## Abstract

Electronic health records provide an opportunity to study prescription drug use in large, heterogeneous populations over time. These data offer a powerful framework for identifying candidate drugs for repositioning against age-related outcomes. In a nationwide cohort of Danish residents aged 50 to 80 years from 2001 to 2015 (*n* = 2,512,380; 23,371,354 person-years), we examined associations between drug exposure at ATC level 4 and 9 major age-related outcomes. Using two complementary analytical approaches—Bayesian time-varying Cox regression and longitudinal minimum loss estimation—we reproduced established drug–outcome associations, including a reduction in 3-year absolute risk of death associated with statins (ATC:C10AA) of −0.8% (95% CI = [− 1.2%, −0.5%]) in females and −0.8% (95% CI = [−1.3%, −0.2%]) in males. Across the analyses, we identified 76 drug–disease pairs warranting further investigation. Among these, biguanides (ATC:P01BB) showed particularly notable associations: the 3-year absolute risk of death was reduced by −1.8% (95% CI = [−2.6%, −1.0%]) in females and −3.9% (95% CI = [−5.0%, −2.7%]) in males. We also observed a potential reduction in cancer risk of −0.3% (95% CI = [−1.9%, −1.3%]) in females and a significant reduction of −1.7% (95% CI = [−3.6%, −0.2%]) in males. Our study provides a general framework for systematically identifying drug–outcome associations in large-scale health registries and prioritizing candidates for further investigation in aging research. While these findings should be interpreted cautiously due to the observational nature of the data, they may help generate hypotheses for future experimental and clinical studies.

## 1 Introduction

The development of new drugs is a time-consuming and expensive process with high rates of attrition often caused by efficacy- and safety-related failures [1–3]. Drug repurposing, re-utilizing approved drugs for indications other than their intended purpose, provides an intriguing proposition due to the reduced risk of adverse side effects given the prior assessment and evaluation for safety and dosing [4]. Repurposing candidates have largely been based on pharmacology and retrospective analysis, with the most notable successes being serendipitous, such as sildenafil for erectile dysfunction [5] or thalidomide for Multiple Myeloma [6] and Acute Myeloid Leukemia (AML) [7].

More systematic studies to produce testable hypotheses range from experimental approaches like binding assays and phenotypic screening to computational methods such as genetic association studies, molecular docking, signature matching, pathway mapping, or the mining of Electronic Health Records (EHRs).

EHRs contribute a rich longitudinal and phenotypic data source, providing real-world evaluations of drug usage in large heterogeneous patient cohorts over prolonged time periods. The era of big data, along with the development of new methods in the context of risk prediction and causality from observational data [8, 9], has opened new opportunities to leverage these data for novel insights [10, 11].

These approaches have already informed clinical decision-making [12–14] and are particularly useful when a randomized clinical trial may not be feasible due to time or ethical constraints. This was especially apparent during the SARS-CoV-2 pandemic, when these methods were used to evaluate the comparative effectiveness of different vaccination and booster campaigns in a rapidly evolving environment [15]. Additionally, conducting an observational study with a target trial in mind can help avoid certain statistical pitfalls such as immortal time bias [16] and provide more robust effect estimates [17].

Here, we make use of the Danish National Prescription Registry (DNPR) [18] and the Danish National Patient Registry (LPR) [19], combining information on all prescribed drugs dispensed through Danish community pharmacies and secondary care diagnoses across Denmark since 1995. We examine the joint contributions of an individual’s drug usage and their comorbidities on the corresponding risk of onset for 9 extensively studied major disease outcomes (dementia, extrapyramidal disorders, coronary vascular disease (CVD), renal failure, chronic obstructive pulmonary disease (COPD), liver disease, inflammatory bowel disease (IBD), cancer, and death). A schematic representation of the study is shown in Figure 1. The first analytic approach is based on a Bayesian version of a time-varying Cox regression model as described previously [20]. This provides an evaluation of the multivariate effect size of the dispensed drug on the specified disease outcomes. As a second analysis, we use longitudinal targeted minimum loss estimation (LTMLE) [21, 22], a doubly robust causal inference method, to obtain robust effect estimates. While we do use causal inference methods, they are applied generically across most drugs and various outcomes, rather than explicitly emulating a specific target trial; therefore, estimates should not be considered causal.

**Figure 1.**
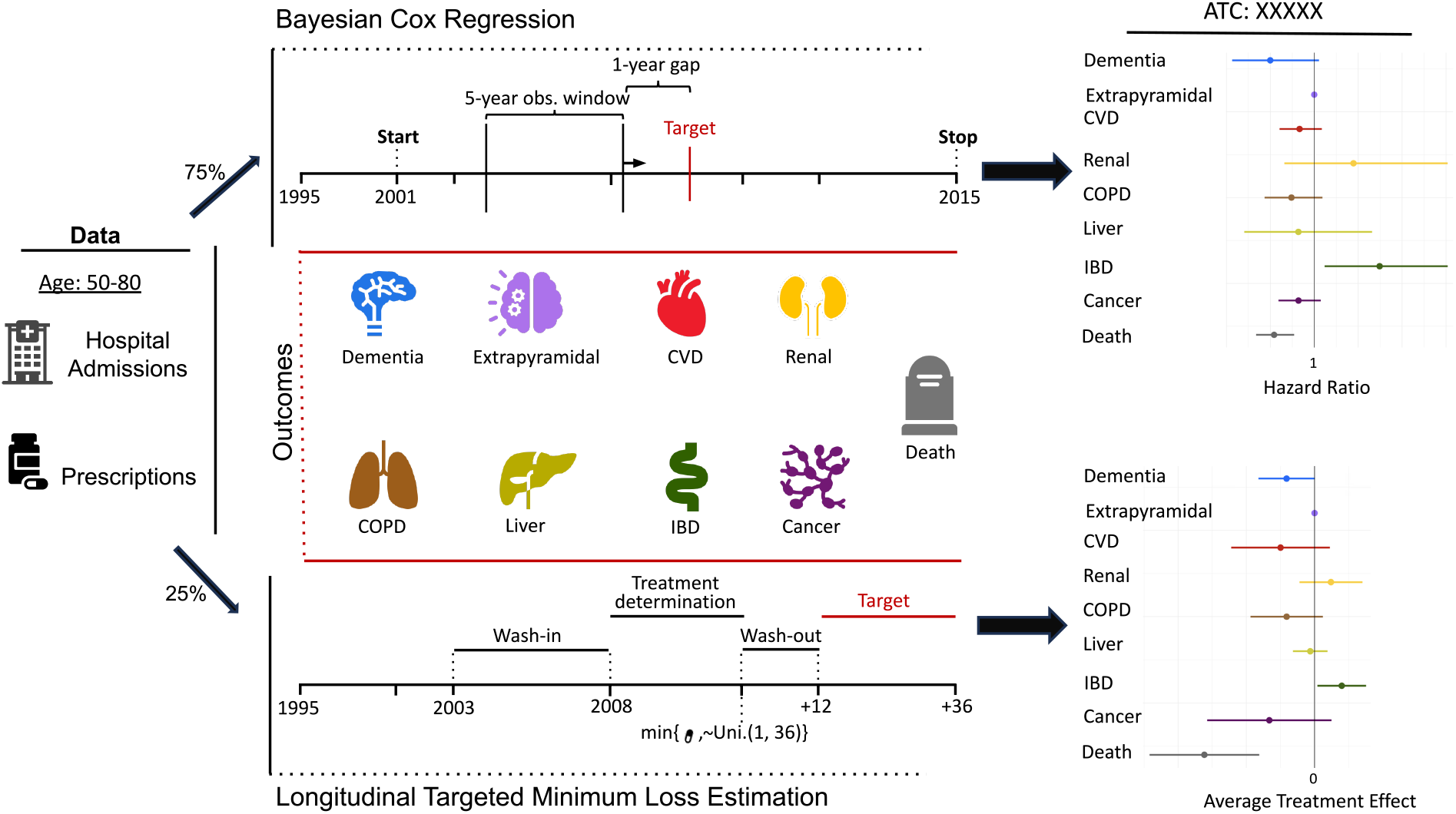
Schematic representation of the study. Information on secondary care admissions and pharmacy-dispensed drugs of individuals residing in Denmark aged 50-80 years is collated. 75% of the individuals are used for a risk prediction study (upper part) and 25% are used for a targeted inference study (lower part). For both approaches, 9 outcomes are evaluated. The final results are effect estimates for a specific drug across the different outcomes, measured as either hazard ratio or average treatment effect for the different study designs, respectively.

This is a hypothesis-generating study that provides a broad screen of dispensed drugs and their effects on selected major disease outcomes, thereby identifying a set of potential repurposing targets that could be corroborated with further evidence in the literature and prioritized for further investigation.

## 2 Results

The study is based on all individuals residing in Denmark aged 50-80 years during a time window from 2001 until 2015 covering more than 2.5 million individuals and a combined 23 million years of observation containing 12 million diagnoses and 44 million dispensed drugs (Supplementary Table 1). At any given point in time, the covariates comprise binary indicators for secondary care diagnoses (ICD-10-3rd level codes e.g. E11 - Type 2 diabetes mellitus, Chapters: I-XVII: 1125/1034 - females/males) and binary indicators for dispensed drugs (ATC-4th level codes, e.g. A10BA, total: 472/458 - females/males) in the past 5 years. Separate models for females and males are estimated to investigate potential differential effects between the sexes.

Overall, we use two study designs. First, we use a classical risk prediction design based on a cross-section of the population (75%), on which we estimate a time-dependent Cox regression model for each of the 9 outcomes as a 1-year ahead prediction (upper section Figure 1). Second, we use an emulated target trial design covering the remaining 25% of the population to obtain robust estimates for all drug-disease pairs (lower section Figure 1). The full protocol for the emulated target trial can be found in the Methods section. In brief, the hypothetical trial for a specific treatment (ATC drug) and outcome starts in 2008. Every individual aged 50-80 who had not yet experienced the outcome and had not received the treatment in the past 5 years is eligible for inclusion. Each individual is assigned a random time between 1 and 36 months. If an individual starts treatment during this time window, they are in the treatment arm; otherwise, they are assigned to the control arm. An individual’s treatment time is either the allocated random time or the time at which they start treatment, whichever comes earlier. An additional 12-month washout period is added to the treatment time to avoid confounding by indication but also to allow for a phase-in time of the drug and to align it with the risk prediction design. It should be noted that this may introduce a selection bias; however, considering disease latency and the fact that these are approved drugs, we considered this the preferable trade-off. Subsequently, this determines the start time of the trial with the next 36 months being used as the observation period on which the counterfactual estimates are based.

An overview of the number of individuals in the two designs as well as some basic characteristics is shown in Table 1 (additional information can be found in Supplementary Table 1). The numbers for the emulated target trial are only approximate because they depend on each treatment/outcome pairing. A table for ATC:C10AA (Statins) and CVD is given in Supplementary Table 2, with all other combinations provided in the Supplementary Data.

**Table 1.**
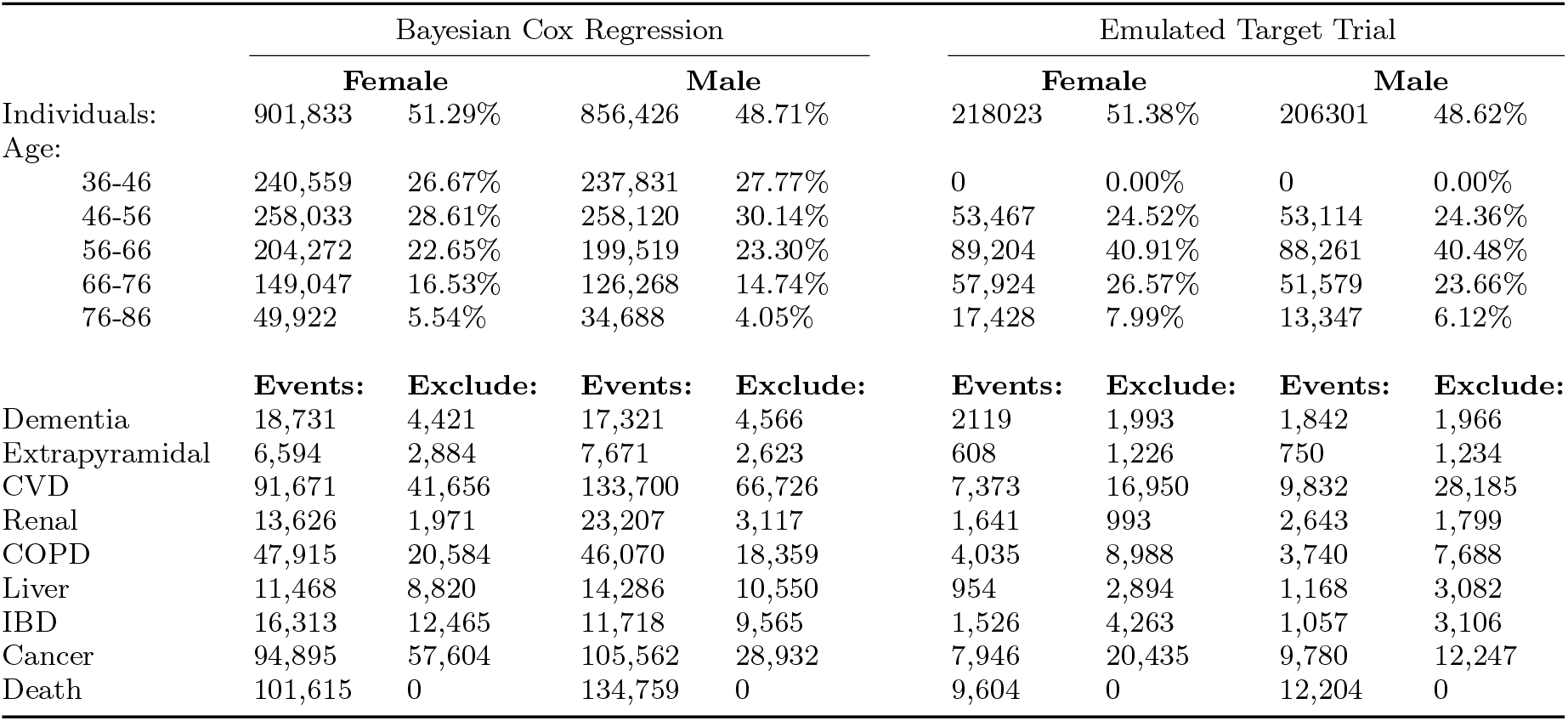
Data overview. Number of individuals in the two study designs split by sex with some basic characteristics. Additionally, the number of events for each outcome is shown as well as the number of excluded individuals due to protocol violations, e.g. prior disease indication. The numbers for the target trial are approximate and should be understood as a rough guide, as each case depends on the treatment/outcome combination.

### Risk assessment of comorbidities and medication history

To understand the overall contribution of secondary care diagnoses and dispensed drugs to the risk of developing one of the 9 outcomes, we conducted a risk prediction study using penalized time-dependent Cox regressions.

Overall, as depicted in Figure 2a and Supplementary Table 3 age-adjusted concordance evaluated on an independent test set demonstrates good discrimination across most of the 9 outcomes with an average concordance of 0.692 (s.d.=0.08) and 0.68 (s.d.=0.079) for females and males, respectively. Cancer exhibits the least predictability with a concordance of 0.552 (95% CI =[0.548, 0.556]) for females and 0.56 (95% CI =[0.556, 0.564]) for males in line with previous studies [23]. Conversely, renal failure and death show the best discrimination with 0.798 (95% CI =[0.79, 0.806]) and 0.798 (95% CI =[0.796, 0.80]) for females and 0.775 (95% CI =[0.769, 0.781]) and 0.774 (95% CI =[0.772, 0.776]) for males. Generally, discrimination between the sexes is similar, with the largest difference being observed for COPD with a concordance of 0.684 (95% CI =[0.68, 0.688]) for females and 0.62 (95% CI =[0.614, 0.626]) for males.

**Figure 2.**
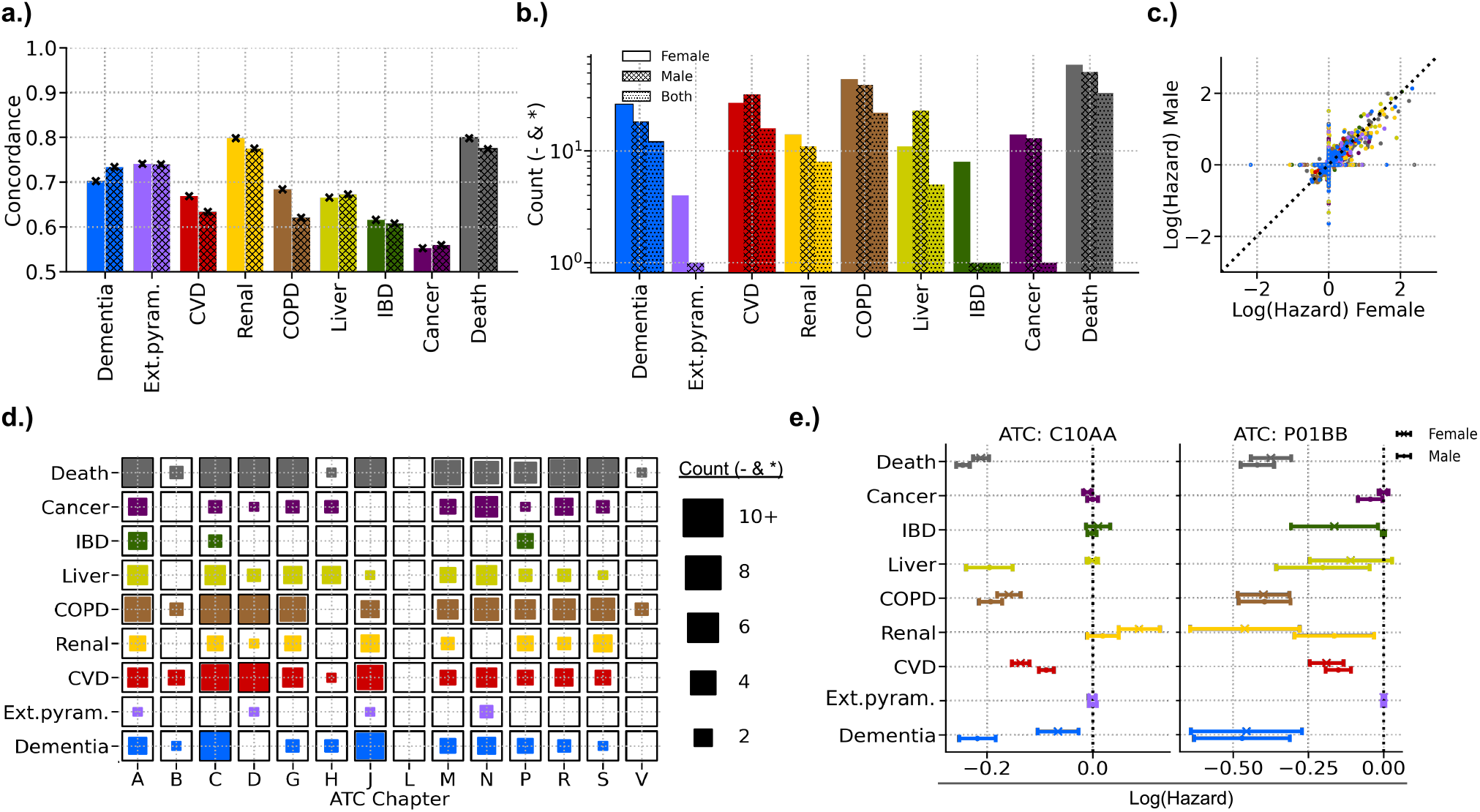
**a**: Age-sex adjusted concordance on test set (25%) across all outcomes. **b**: Number of negative and significant effects (based on highest posterior density 95%) across all outcomes split by sex and combined (counting effects that are present in both sexes). **c**: Scatter plot of the log(hazard) estimates between the model for females and males colored by the corresponding outcome that is estimated. **d**: Number of negative and significant associations aggregated by ATC chapter for each outcome. Effects for females and males are combined and treated as individual estimates. **A** ALIMENTARY TRACT AND METABOLISM, **B** BLOOD AND BLOOD FORMING ORGANS, **C** CARDIOVASCULAR SYSTEM, **D** DERMATOLOGICALS, **G** GENITO URINARY SYSTEM AND SEX HORMONES, **H** SYSTEMIC HORMONAL PREPARATIONS, EXCL. SEX HORMONES AND INSULINS, **J** ANTIINFECTIVES FOR SYSTEMIC USE, **L** ANTINEOPLASTIC AND IMMUNOMODULATING AGENTS, **M** MUSCULO-SKELETAL SYSTEM, **N** NERVOUS SYSTEM, **P** ANTIPARASITIC PRODUCTS, INSECTICIDES AND REPELLENTS, **R** RESPIRATORY SYSTEM, **S** SENSORY ORGANS, **V** VARIOUS. **e**: Forest plot of the effect estimates for ATC:C10AA (Statins) and ATC:P01BB (Biguanides) across all outcomes and split by sex.

As the aim of this study is to identify potential candidates for repurposing, we focus on significant negative effect estimates based on the highest posterior density of 95% (HPD) [Figure 2b]. For females, there are a total of 207 drugs associated with the 9 outcomes. Extrapyramidal disorders show the fewest associations with only 4, while death has the most associations with 59. Similarly, for males, a total of 189 drugs are associated with the outcomes, with the fewest associations identified for extrapyramidal disorders and IBD, each with only 1 drug, and the most associations identified for death, with 51 drugs. Comparing across sexes, we identify a total of 98 drugs that show an effect in both sexes, with extrapyramidal disorders having none, IBD and cancer having only 1 each, while death has the most with 33 common associations. Forest plots for all significant estimates, irrespective of the direction of the effect for each outcome, can be found in Supplementary Figures 1-9. The entire set of estimates can be found in the Supplementary Data.

Overall, effect estimates largely agree across sexes, showing high degrees of correlation between the log(hazard) estimates, as depicted in Figure 2c. All outcomes show Pearson correlations above 0.6, except for cancer, which shows a correlation of 0.459 [Supplementary Table 4]. Visual inspection of Figure 2c reveals that most estimates lie on the diagonal, indicating good agreement. However, some points lie on the respective axes, indicating estimates close to 0 for either sex. This does not necessarily reflect a true effect size of 0 but might instead be a result of the penalization term and a corresponding lack of power.

Furthermore, summarizing the effects within the corresponding ATC chapters in Figure 2d reveals overall patterns of drug-disease pairs. For instance, drugs in the chapter Alimentary tract and Metabolism (A) show an effect for all 9 outcomes, followed by drugs categorized in the cardiovascular system (C) and drugs in antiparasitic products, insecticides, and repellents (P), both missing associations with extrapyramidal disorders only. No negative associations are found for drugs in the chapter antineoplastic and immunomodulating agents (L).

However, the most meaningful insights can be gained when examining individual drugs across all outcomes simultaneously, as exemplified in the case of ATC:C10AA (Statins) and ATC:P01BB (Biguanides) in Figure 2e. Consistent with published clinical trial results [24, 25], ATC:C10AA shows a reduced risk of death, with a log(hazard) of -0.211 (95% HPD =[-0.226, -0.196]) for females and -0.246 (95% HPD =[-0.259, -0.233]) for males, as well as a reduced risk of CVD, with a log(hazard) of -0.136 (95% HPD =[-0.153, -0.12]) for females and -0.088 (95% HPD =[-0.102, -0.074]) for males.

Surprisingly, ATC:P01BB, a biguanide used for the treatment and prevention of malaria, shows clear negative associations across most of the evaluated disease outcomes, with similar patterns across the sexes. However, this could be due to unmeasured confounding, as people who use anti-malaria drugs might be traveling and hence are likely in an overall healthy state. On the other hand, Biguanides classified in ATC:A10AB, used in diabetes care, e.g. metformin, show either no effect or an increase in risk [Supplementary Figure 10].

### Targeted evaluation of drug-disease pairs

To gain further insight into the reliability of the estimates, we perform a targeted inference analysis across most drug-disease pairs (only combinations with at least 1,000 treated individuals with 100 events). As mentioned earlier, this analysis is conducted in a generic way to scale to the number of combinations analyzed here, a total of 890 for females and 742 for males, rather than through carefully tailored target trials; therefore, results should be interpreted cautiously.

In total, we identify 76 drugs that show a significant negative association with the outcomes, with 52 for females, 24 for males, and 12 common across sexes. Dementia and death exhibit the most associations, with 21 and 10 for females, and 4 and 10 for males, respectively. Cancer and IBD show the fewest associations for females, with only 1 drug identified for each, whereas IBD shows no effects for males. Common associations between sexes are only identified for dementia, CVD, COPD, and death [Figure 3a].

**Figure 3 a:**
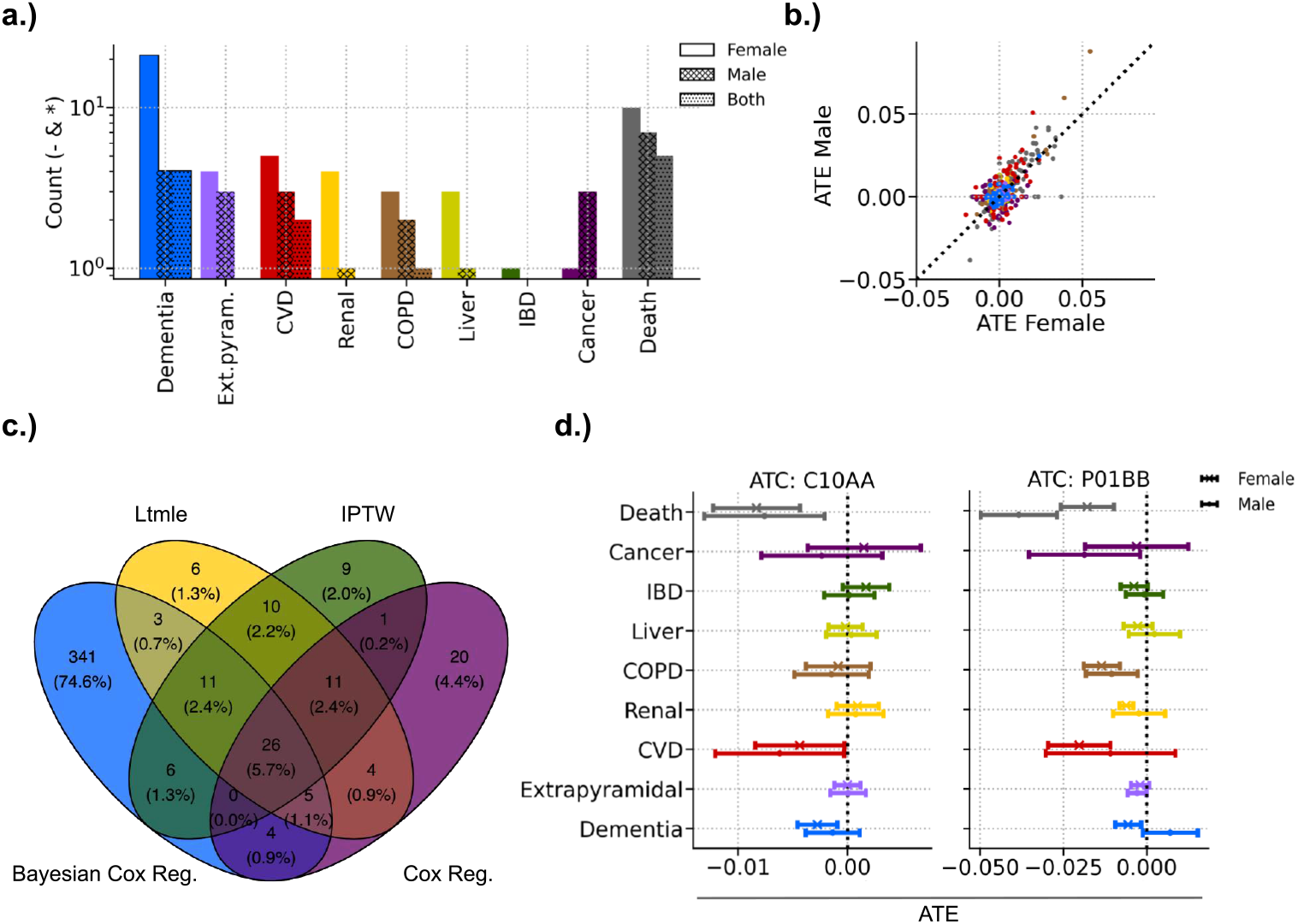
Number of negative and significant effects (based on the 95% confidence interval) across all outcomes split by sex and combined (counting effects that are present in both sexes). **b**: Scatter plot of the average treatment effect (ATE) estimates between the model for females and males colored by the corresponding outcome. **c**: Venn diagram of the count of overlapping negative and significant effects between the risk prediction study and estimates based on the emulated target trial, including longitudinal targeted minimum loss estimation (LTMLE), inverse probability of treatment weighted estimation (IPTW), and a simple Cox regression. **d**: Forest plot of the effect estimates for ATC:C10AA (Statins) and ATC:P01BB (Biguanides) across all outcomes and split by sex.

Overall, estimates for females and males appear similar, as can be seen in Figure 3b, with most estimates being close to the diagonal. This similarity is also reflected in the correlation of the average treatment effect (ATE) estimates between the sexes, with all outcomes showing a correlation of at least 0.2, except for cancer, which has a correlation of 0.129 (Supplementary Table 5).

Furthermore, estimates are relatively stable between the two study designs and across different approaches, as illustrated in Figure 3c. Here, we present a Venn diagram of the identified significant negative associations for drug-disease pairs aggregated over the sexes. We compare the estimates from the Bayesian Cox regressions with the LTMLE estimates, inverse probability of treatment weighting estimates (IPTW), and estimates from simple Cox regressions fitted to the emulated trial data. A total of 45 (9.9%) estimates appear similar between the Bayesian Cox regression and LTMLE. A large fraction of 341 (74.6%) associations are only identified in the Bayesian Cox regressions; however, this is also expected as this design has the most power and is potentially more prone to identifying spurious relations. Estimates largely overlap with a total of 26 (5.7%) associations identified in all approaches. It should be stressed here that the estimates from the Bayesian Cox regression and LTMLE are inherently different estimands: one is a relative risk and the other is a counterfactual absolute risk. Only LTMLE with IPTW and the two Cox regressions can be directly compared with each other. Hence, the depiction should be understood as a high-level summary rather than a direct comparison. A table showing all estimates across the approaches can be found in the Supplementary Data.

Lastly, we again examine the effects of individual drugs across all outcomes simultaneously, as exemplified here by ATC:C10AA (Statins) and ATC:P01BB (Biguanides) in Figure 3d. Several of the effects identified for ATC:C10AA from the risk prediction study vanish, with the main known effects from clinical trials remaining significant, albeit slightly attenuated. Death shows an absolute risk reduction of -0.8% (95% CI =[-1.2%, -0.5%]) and -0.8% (95% CI =[-1.3%, -0.2%]) over a 3-year period for females and males, respectively, while CVD indicates a reduced absolute risk of -0.5% (95% CI =[-0.8%, -0.0%]) and -0.6% (95% CI =[-1.2%, -0.0%]).

Interestingly, we still observe a significant effect for ATC:C10AA and dementia in females, with a reduced absolute risk of -0.3% (95% CI =[-0.5%, -0.1%]). ATC:C10AA also shows a negative effect for males, with an absolute risk reduction of -0.2% (95% CI =[-0.4%, 0.1%]), although there is no clear evidence for the direction of the effect. While a potential link between statins and dementia risk has been proposed earlier [26–28], evidence to date is inconclusive [29] and further investigation may be warranted.

Surprisingly, many of the effects identified in the risk prediction study for ATC:P01BB persist. In total, we find 8 significant effects for ATC:P01BB across all outcomes and both sexes. Death shows a clear 3-year absolute risk reduction of -1.8% (95% CI =[-2.6%, -1.0%]) and -3.9% (95% CI =[-5.0%, -2.7%]) for females and males, respectively. Further, we identify a potential absolute risk reduction for cancers of -0.3% (95% CI =[-1.9%, 1.3%]) in females and a significant reduction in absolute risk of -1.7% (95% CI =[-3.6%, -0.2%]) for males. Most of the effects are similar between the sexes, albeit with stronger evidence of a sign effect in females. Dementia is the only exception to this, with a significant absolute risk reduction of -0.6% (95% CI =[-1.0%, -0.2%]) in females but a potential absolute risk increase of 0.7% (95% CI =[-0.1%, 1.5%]) in males.

Looking at ATC:A10AB in Supplementary Figure 10, a different Biguanide containing Metformin, we mostly see no clear effects, with the exception of death in males with an absolute risk increase of 1.6% (95% CI =[0.2%, 2.9%]) and liver disease in males with an absolute risk increase of 0.6% (95% CI =[0.1%, 1.2%]).

While the effects identified for ATC:P01BB appear interesting as a potential drug class for further investigation, as we have cautioned in the previous section, there are potential mechanisms of confounding that we cannot control for, which could have biased the effects toward a reduction in risk. A recent preprint has reported results congruent with our findings [30]. This study is based on data from an Israeli healthcare organization, but importantly conducts validation in a large international collection of EHR data using active comparators rather than non-treated individuals.

The purpose of the study is solely hypothesis-generating, and hence the effects need to be verified in other, more targeted studies and further corroborated through additional evidence. A minimal next step would be to investigate biomedical databases for potential links. As an example, we looked at links between drugs in ATC:P01BB and our disease outcomes in the precision medicine database PrimeKG [31]. Traversing the knowledge graph from associations between drugs and genes/proteins, annotated as carrier, enzyme, target, or transporter, and subsequently from genes/proteins to our disease outcomes, as shown in Figure 4, reveals a possible relation to several of the outcomes, with Chloroquine as a particularly outstanding case. Another possible mechanism of action reported in the literature is the link to the use of Hydroxychloroquine in the treatment of rheumatoid arthritis and other inflammatory rheumatic diseases [32]. Generally, we do see several anti-inflammatory drugs, e.g. Corticosteroids (ATC: C05AA, D07AB, H02AB, R01AD) or Anti-inflammatory and anti-rheumatic agents, non-steroids (ATC: M01AB, M01AX) with potential repositioning effects, possibly indicating underlying inflammatory aspects to some of the disease outcomes (Supplementary Figure 11). Further, recent studies suggest potential risk-reducing effects of Hydroxychloroquine on dementia risk [33] or atovaquone in the treatment of non-small cell lung cancer [34].

**Figure 4.**
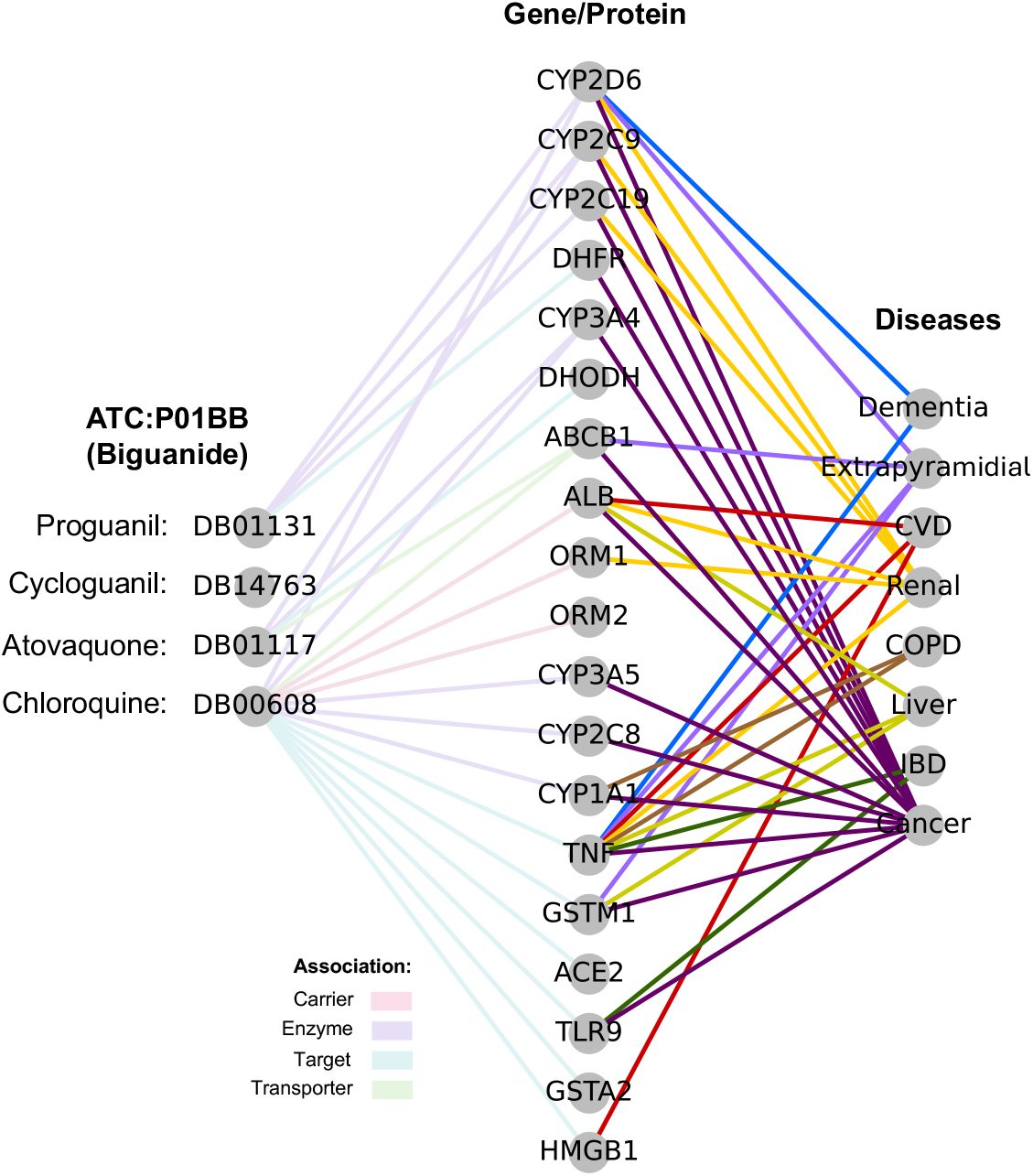
PrimeKG knowledge graph extract for drugs in ATC:P01BB (Biguanides) and their association with genes/proteins and their respective associations with the disease outcomes.

## 3 Discussion

Overall, this study demonstrates that EHRs may be helpful in identifying novel associations that warrant further investigation. We conducted a comprehensive screening of most ATC level-4 drugs across a range of disease outcomes. All estimates are provided and can serve as a foundation for subsequent research or as supplementary evidence to support findings obtained from alternative approaches.

The most effective way to utilize these types of data and methods is through a carefully crafted emulated target trial with proper inclusion criteria. However, this is usually only possible with a specific hypothesis in mind. While our approach can easily be applied generically across most combinations, this also makes it more prone to misspecification and may introduce biases (e.g. selection bias).

Other limitations of our approach include the possibility of confounding from various sources. While we do control for a comprehensive set of medical information on individuals, there are certain aspects that may not be adequately addressed. Improved access to a broader range of clinical data could potentially mitigate some of these issues; however, the effects should be assessed with appropriate domain expertise to identify potential biases and directions of influence. Furthermore, information on medications includes only prescriptions and does not necessarily reflect actual use. These two sources could explain the effects found for ATC:P01BB. First, it is not clear if individuals are actually taking the drug or only received a prescription due to planned travel. Second, individuals who go on long-distance travel are probably healthier and potentially socioeconomically better off, which we can only partially control for.

As mentioned earlier, the washout period may introduce a selection bias. While in this study we argue that the impact of confounding by indication is potentially more critical, motivating our choice, better information on clinical and socioeconomic factors could help mitigate this effect and allow for better designs.

Furthermore, our design results in a real-time shift between the treatment and control arms, where treatment consistently occurs before the allocated random time. This shift should not significantly impact effect estimates unless there is a noticeable change in incidence within a small time window. However, during rapid shifts in disease incidence, such as those observed during the SARS-CoV-2 pandemic, this could become important and should be taken into consideration. We conducted evaluations of the potentially introduced bias through simulations (see supplementary data) and found no measurable effect.

A potential extension to our approach could involve incorporating multiple time points for the emulated trial, thereby enhancing the overall power of the approach. Additionally, this approach would enable the study of outcomes that are typically rare, potentially offering more opportunities for drug repositioning.

Finally, our approach assesses a wide array of combinations. While we use shrinkage priors and incorporate multiple cohort splits (such as data and sex splits), thereby offering multiple lines of evidence, we do not adjust for multiple hypotheses. It is important to emphasize that the objective of the study is not to make inferential statements about a particular effect, but rather to explore and screen for new targets.

## 4 Methods

### Risk prediction

#### Data sources

This study retrospectively uses data from the Danish health registries, which include the Central Person Registry (CPR), the Danish National Patient Registry (LPR), the Death Registry (DR), and the Danish National Prescription Registry (DNPR). Individuals born in or residing in Denmark for more than 3 months are registered. All registries have been linked via a unique personal identifier. Data compilation spans from January 1, 1995, to December 31, 2014.

#### Cohort

We included all individuals aged 50 to 80 who were alive on January 1, 2001, and who had been residing in Denmark continuously since at least January 1, 1995. This inclusion criterion ensures that all participants have a minimum of five years of recorded medical information at any given point in time. Participants exit the cohort upon reaching the age of 80 or due to exclusion criteria such as emigration, end of follow-up or death, whichever occurred first. Individuals who emigrated after January 1, 2001, are censored at the point of emigration and remain so for the duration of the study. Individuals with events of interest occurring prior to January 1, 2001, are excluded from the study. The primary observational period for model fitting and evaluation extends from January 1, 2001, to December 31, 2014. The cohort is divided into three subsets: (i) a training set (70%), used for model training and development; (ii) a validation set (5%), used for initial model evaluation and to determine the optimal penalization strength; and (iii) a test set (25%), used for the final model assessment.

#### Covariates

The covariates include binary indicators for secondary care diagnoses extracted from the Danish National Patient Registry (LPR), utilizing ICD-10 codes up to the third level of specificity, recorded across chapters I-XVII (e.g., E11 for Type 2 diabetes mellitus). Considering the sex specificity of some diagnoses, we filtered indicators relevant to each sex, resulting in a total of 1,125 indicators for females and 1,034 indicators for males. We limited the indicators to records from the preceding five years at any given point in time to capture recent health changes. Similarly, binary indicators for dispensed medications, recorded in the Danish National Prescription Registry (DNPR), are included. These medications are classified using the ATC system at the fourth level (e.g., A10BA for Biguanides). Because some medications are sex-specific, we identified a total of 472 indicators for females and 458 for males, respectively. These indicators reflect medication usage in the past five years.

#### Outcomes

We consider a total of 9 outcomes including dementia (ICD10: F00-03, G30-31), extrapyramidal disorders (ICD-10: G20-26), coronary vascular disease (CVD) (ICD-10: I21-26, I46, I50, I60-64), renal failure (ICD-10: N17-19), chronic obstructive pulmonary disease (COPD) (ICD-10: J41-J44, J47), liver disease (ICD-10: K70-77), inflammatory bowel disease (IBD) (ICD-10: K50-52), cancer (ICD-10: C00-96, D37-48 excluding: C44, D45), and death.

#### Statistical analysis

We fit time-dependent Bayesian Cox models with shrinkage priors for each of the nine outcomes and for each sex, using age as the underlying timeline. These models are solely fitted on the training set. Individuals are considered at risk upon reaching the inclusion age or the age at which they enter the cohort, whichever comes first. They are followed until the occurrence of a specific outcome, death, emigration, or the end of the follow-up period. Covariates are treated as time-dependent, consisting of binary indicators for diseases and medications within the preceding five years. The effects of these covariates are modeled through a linear predictor. To prevent the inclusion of data that may only reflect the diagnostic process leading up to an outcome, we introduce a one-year gap between the occurrence of an event and its associated covariates. This approach ensures that evaluations are based on predictions made at least one year in advance (making it similar to the LTMLE estimate with a 1-year washout).

Technically, we are interested in deriving the posterior distribution *p*(***θ***|**D**) ∝ ℒ (**D**|***θ***)*p*(***θ***) of the model parameters ***θ*** conditioned on the observed data **D** = {(*y*_*i*_ := min{*T*_*i*_, *u*_*i*_}, *δ*_*i*_, **X**_*i*·_(*t*)) : *i* = 1, 2, …, *n*} for *n* individuals, where *T*_*i*_ is the event time, *u*_*i*_ is the censoring time, *δ*_*i*_ is an indicator for censoring, and **X**_*i*·_(*t*) corresponds to the p-covariate processes **X**_*i*·_(*t*) = (*X*_*i*1_(*t*), …, *X*_*i*p_(*t*))^⊤^. We follow Jung et al. (2022, 2023) [20, 23] and use the reweighted Cox partial log-likelihood based on a counting process representation,

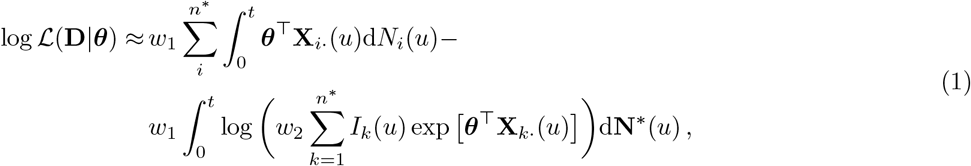

where 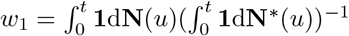 i.e., the ratio of events, *w* = *n*(*n*^∗^)^−1^ i.e., the ratio of observations, *I*_i_ (*t*) as the risk set indicator of individual *i* at time *t*, with **N** representing the corresponding multivariate counting process ∗ with denoting potentially subsampled cohorts during inference. For the final model, we use as prior distribution *p*(***θ***) a Student-T distribution,

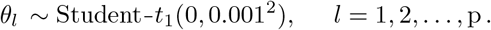

Inference is done through variational methods with a low-rank (50) multivariate normal distribution as the distributional family for stochastic variational inference. We perform stochastic gradient descent updates using batches of 8,196 randomly selected individuals. Credible regions or highest posterior density intervals (HPD) are determined based on the posterior distributions, typically covering a 95% credible interval unless stated otherwise. The concordance index serves as the primary metric for evaluating the model fits. For a more detailed description of the method and implementation, we refer to the aforementioned articles.

While our primary interest lies in the hazard ratios, it should be noted that the hazard ratio is not valid as a causal estimand in general, as the different risk sets under varying regimes are not comparable if there is a nonnull effect of treatment [35, 36]; hence, it cannot directly be compared to the estimates from the LTMLE analysis. Additionally, the hazard ratio only provides an estimate for the relative risk. In order to obtain a similar absolute risk estimate, one can use Breslow’s estimator [37] to obtain an estimator for the baseline hazard 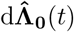 and then obtain the absolute risk estimate over a time period from *l* to *t* as 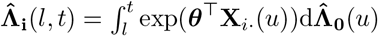.

However, we refrain from estimating the absolute risk here as the estimation of the baseline hazard is computationally intensive and our main interest is on whether there is a directional effect rather than its proper quantification.

### Targeted inference

#### Data sources

We use the same dataset as in the observational study, specifically all individuals from the test set. As the test set was only used to evaluate the concordance index for the observational study, it constitutes an independent data subset for estimation purposes. In principle, all individuals from the test set are included; however, additional restrictions will apply based on the targeted trial design, which we address below.

#### Covariates

The covariates used for estimation mirror those used in the observational study. However, instead of treating them as time-dependent variables, we focus on a single time point: the start time of the emulated trial, excluding the washout period. From this time point, we construct binary indicators representing medication usage and acquired diseases over the past five years. Additionally, we apply a filtering criterion to the covariates, ensuring a minimum frequency of 0.01 in the entire population, the treatment group, the untreated group, the event group, the non-event group, or any combination thereof, for each treatment-outcome pairing separately. This step aims to eliminate covariates that occur in only a small fraction of individuals across all possible subgroups, thereby reducing computational burden. Furthermore, we add indicators for age at trial start in 5-year brackets from 50 to 75.

#### Treatments

We consider all ATC level-4 drugs as potential treatments as this level of granularity provides the best trade-off in our data between specificity of the drugs used and reasonably sized treatment groups. However, we restrict our analysis to drugs with a minimum of 1,000 treated individuals in a given emulated target trial.

#### Outcomes

Same as for the observational study.

#### Eligibility criteria

Eligibility criteria are specific for each treatment and outcome. The start date for each trial is January 1, 2008. Individuals have to be between 50 and 80 years of age to be able to join. Furthermore, the specific outcome under study should not have occurred prior to the start date. Individuals who had an indication of treatment in the preceding 5 years (January 1, 2003) are excluded.

#### Treatment assignment

All individuals eligible for the trial on January 1, 2008, are assigned a random time, uniformly drawn from 1 to 36 months. If an individual has an indication of treatment within this time window, the earliest time of treatment initiation is set as the new allocated time, and the individual enters the treatment arm. If no treatment indication is registered in the time window, the individual enters the control arm. Individuals in the treatment arm are assumed to stay on treatment during the entire study period. We do not consider treatment discontinuation as it is difficult to define generically valid intervals of treatment interruption. Individuals in the control arm who subsequently switch to treatment are censored after a washout period of one year after the switch.

#### Treatment strategies

The strategies to be compared are (i) initiation of treatment and presumed continuation over the study period and (ii) no initiation of treatment during the entire study period.

#### Follow-up

After treatment determination, every individual goes through a 1-year washout period to avoid treatment-by-indication effects and to allow for a phase-in period of the drug. If an event occurred during treatment determination or the washout period, individuals are removed from the study. The time after the washout period constitutes time 0 and the start of the trial. All remaining individuals are followed for a maximum of 36 months, until occurrence of the event, or until possible censoring (death or emigration).

#### Causal estimands

The primary outcome of the study is the average treatment effect between the treatment group and the control group after 36 months, measured as the difference between the absolute risk in the two arms, i.e.

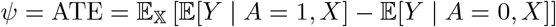

where *Y* is the absolute risk after 36 months, *A* is the treatment indicator, and *X* is a vector of covariates.

#### Statistical analysis

The primary statistical method for the emulated target trial analysis is based on a targeted minimum loss estimator (TMLE) for the parameters of longitudinal static and dynamic marginal structural models as implemented in R-ltmle [22]. The observed data for an individual *i* are *O*_*i*_ = (*L*(0), *A*(0), *Y* (1), *C*(1), …, *A*(*K*), *Y* (*K*), *C*(*K*)) for *K* discrete time points, with *L*(0) as the baseline covariates, *A*(*t*) as the intervention node describing the treatment status, *C*(*t*) the censoring status, and *Y* (*t*) as the outcome process at time *t*. We observe for *n* individuals **O** ∼ *P*_*O*_ with *P*_*O*_ ∈ 𝒫, the set of all possible distributions that could give rise to **O**. We assume that *P*_*O*_ can be factorized as

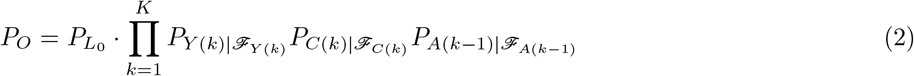

where ℱ_∗(*k*)_ describes the filtration of node ∗ up to *k*. Under this specification 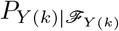 can be understood as the g-formula/outcome model and 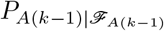 as the propensity score.

We are interested in a static treatment regime comparing continuous treatment *d*_1_ with no treatment *d*_0_, in a hypothetical no-censoring scenario, e.g. *d*_1_(*k*) = (*A*_0_ = 1, …*A*_*k*_ = 1, *C*_1_ = 0, *C*_*k*_ = 0)). Our target parameter of interest is then 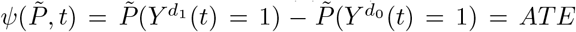, where 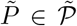 is the distribution for the hypothetical counterfactual population.

Under the assumptions of sequential exchangeability, consistency, and positivity, we can identify the target parameter from the observed data and estimate 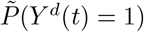 via a sequence of nested expectations [21].

For our implementation, we split time into 6-month intervals. Each sequential fit is performed via a parametric generalized linear model. Overall, the default parameters for R-ltmle are used. We extract the TMLE estimate for our primary end point of the 36-month absolute risk between the treatment and control group, plus the corresponding 95% confidence interval. Similarly, we extract the corresponding IPTW estimator from the same procedure (automatically estimated for TMLE). Finally, we also fitted a simple Cox regression on the emulated target trial data with the covariates and a treatment indicator as an additional comparator.

#### Causal interpretation

Causal interpretation of the results from the Cox regression analyses and the target trial emulation is hampered by the fact that the identification strategy is naive. As highlighted in our discussion, there is likely considerable unobserved confounding of the results of both sets of analyses. Additionally, our generic design for the target trial emulation, while easy to scale to many treatment-outcome combinations, is limited.

## Data Availability

Danish registry data are available for use in
secure, dedicated environments via application to the Danish Patient Safety Authority and
the Danish Health Data Authority via
https://sundhedsdatastyrelsen.dk/da/english/health_data_and_registers/research_services/a
pply.

## Contributors

AWJ developed the methods and the study design, conducted the analysis, assembled all figures, and wrote the manuscript. IL and SB provided overall feedback and guidance, as well as help with the drug databases. AWJ and LHM conceived the study and accessed and verified the Danish data. LHM and SB supervised the study. All authors had full access to all the data in the study and had final responsibility for the decision to submit for publication.

## Declarations of interests

SB received personal compensation for managing board memberships at Intomics and Proscion and is a scientific advisory board member of Biocenter Finland, Health Data Research UK, the Finnish Center of Excellence in Complex Disease Genetics, ELIXIR Node (Luxembourg), Lund University Diabetes Centre (Lund, Sweden), and SciLifeLab (Stockholm, Sweden). SB reports stock holdings in Intomics, Hoba Therapeutics Aps, Novo Nordisk, Eli Lilly, and Lundbeck. All other authors declare no competing interests.

## Data sharing

Danish registry data are available for use in secure, dedicated environments via application to the Danish Patient Safety Authority and the Danish Health Data Authority.

Code is available on https://github.com/alexwjung/DrugTarget

## Acknowledgements

This work was supported by the Novo Nordisk Foundation under grants NNF17OC0027594 and NNF14CC0001.

## Notes

### Funding Statement

This work was supported the Novo Nordisk Foundation under grants NNF17OC0027594 and NNF14CC0001.

### Author Declarations

The use of the Danish National Registries was conducted under the Danish Data Protection Act. Furthermore, the analysis was conducted under the information security and data confidentiality policies of Statistics Denmark, which is the Danish National Statistical Institution.

### Summary of Updates

Updated scope of the paper and changed text to better reflect target audience.

