## Supplementary for "Identifying drug repositioning candidates for age-related outcomes in the Danish health registries"

### 1 Supplementary Tables

**Supplementary Table 1** Data overview for the observational design, split into training, validation, and test sets.

| | Train | | | | Valid | | | | Test | | | |
|---|---|---|---|---|---|---|---|---|---|---|---|---|
| | Female | | Male | | Female | | Male | | Female | | Male | |
| Patients: | 901,833 | 51.29% | 856,426 | 48.71% | 64,501 | 51.28% | 61,291 | 48.72% | 322,092 | 51.26% | 306,237 | 48.74% |
| Age: | | | | | | | | | | | | |
| 36-46 | 240,559 | 26.67% | 237,831 | 27.77% | 17,092 | 26.50% | 17,162 | 28.00% | 85,484 | 26.54% | 85,127 | 27.80% |
| 46-56 | 258,033 | 28.61% | 258,120 | 30.14% | 18,420 | 28.56% | 18,260 | 29.79% | 92,020 | 28.57% | 92,593 | 30.24% |
| 56-66 | 204,272 | 22.65% | 199,519 | 23.30% | 14,633 | 22.69% | 14,276 | 23.29% | 73,081 | 22.69% | 71,052 | 23.20% |
| 66-76 | 149,047 | 16.53% | 126,268 | 14.74% | 10,690 | 16.57% | 9,199 | 15.01% | 53,572 | 16.63% | 45,160 | 14.75% |
| 76-86 | 49,922 | 5.54% | 34,688 | 4.05% | 3,666 | 5.68% | 2,394 | 3.91% | 17,935 | 5.57% | 12,305 | 4.02% |
| Obs. Period: | 8,437,523 | 1 - 11 - 14 | 7,920,855 | 1 - 11 - 14 | 602,705 | 1 - 11 - 14 | 563,748 | 1 - 10 - 14 | 3,015,650 | 1 - 11 - 14 | 2,830,873 | 1 - 10 - 14 |
| Unique Diag. | 4,621,418 | 0 - 4 - 16 | 4,276,183 | 0 - 3 - 16 | 330,484 | 0 - 4 - 16 | 306,397 | 0 - 3 - 16 | 1,647,576 | 0 - 4 - 16 | 1,525,370 | 0 - 3 - 16 |
| Unique Pres. | 17,871,825 | 4 - 17 - 44 | 13,093,221 | 2 - 13 - 36 | 1,277,595 | 4 - 18 - 44 | 936,692 | 2 - 13 - 36 | 6,378,036 | 4 - 17 - 44 | 4,679,747 | 2 - 13 - 36 |
| Events: | | | | | | | | | | | | |
| Dementia | 18,731 | 2.08% | 17,321 | 2.02% | 1,303 | 2.02% | 1,263 | 2.06% | 6,743 | 2.09% | 6,279 | 2.05% |
| Extrapyramidal | 6,594 | 0.73% | 7,671 | 0.90% | 455 | 0.71% | 573 | 0.93% | 2,344 | 0.73% | 2,779 | 0.91% |
| CVD | 91,671 | 10.16% | 133,700 | 15.61% | 6,635 | 10.29% | 9,580 | 15.63% | 32,818 | 10.19% | 47,506 | 15.51% |
| Renal | 13,626 | 1.51% | 23,207 | 2.71% | 984 | 1.53% | 1,684 | 2.75% | 4,789 | 1.49% | 8,317 | 2.72% |
| COPD | 47,915 | 5.31% | 46,070 | 5.38% | 3,463 | 5.37% | 3,265 | 5.33% | 17,166 | 5.33% | 16,412 | 5.36% |
| Liver | 11,468 | 1.27% | 14,286 | 1.67% | 799 | 1.24% | 1,070 | 1.75% | 4,114 | 1.28% | 5,098 | 1.66% |
| IBD | 16,313 | 1.81% | 11,718 | 1.37% | 1,204 | 1.87% | 784 | 1.28% | 5,953 | 1.85% | 4,143 | 1.35% |
| Cancer | 94,895 | 10.52% | 105,562 | 12.33% | 6,884 | 10.67% | 7,571 | 12.35% | 34,201 | 10.62% | 37,771 | 12.33% |
| Death | 101,615 | 11.27% | 134,759 | 15.74% | 7,192 | 11.15% | 9,813 | 16.01% | 36,165 | 11.23% | 48,157 | 15.73% |
| Excluded: | | | | | | | | | | | | |
| Dementia | 4,421 | 0.49% | 4,566 | 0.53% | 331 | 0.51% | 308 | 0.50% | 1,673 | 0.52% | 1,655 | 0.54% |
| Extrapyramidal | 2,884 | 0.32% | 2,623 | 0.31% | 212 | 0.33% | 204 | 0.33% | 1,062 | 0.33% | 994 | 0.32% |
| CVD | 41,656 | 4.62% | 66,726 | 7.79% | 3,028 | 4.69% | 4,813 | 7.85% | 14,912 | 4.63% | 23,643 | 7.72% |
| Renal | 1,971 | 0.22% | 3,117 | 0.36% | 137 | 0.21% | 217 | 0.35% | 652 | 0.20% | 1,138 | 0.37% |
| COPD | 20,584 | 2.28% | 18,359 | 2.14% | 1,429 | 2.22% | 1,311 | 2.14% | 7,388 | 2.29% | 6,460 | 2.11% |
| Liver | 8,820 | 0.98% | 10,550 | 1.23% | 578 | 0.90% | 717 | 1.17% | 3,180 | 0.99% | 3,680 | 1.20% |
| IBD | 12,465 | 1.38% | 9,565 | 1.12% | 903 | 1.40% | 653 | 1.07% | 4,453 | 1.38% | 3,322 | 1.08% |
| Cancer | 57,604 | 6.39% | 28,932 | 3.38% | 4,143 | 6.42% | 2,156 | 3.52% | 20,599 | 6.40% | 10,358 | 3.38% |
| Death | 0 | 0.00% | 0 | 0.00% | 0 | 0.00% | 0 | 0.00% | 0 | 0.00% | 0 | 0.00% |

**Supplementary Table 2** Data overview for the target trial design of ATC:C10AA (statins) and CVD.

| **Female** | | | | **Male** | | | |
|---|---|---|---|---|---|---|---|
| | Level | treat_1 = 0 (n=131608) | treat_1 = 1 (n=8691) | Variable | Level | treat_1 = 0 (n=131608) | treat_1 = 1 (n=8691) |
| Age_50_55 | 0 | 111,761 (74.8) | 8,946 (85.2) | Age_50_55 | 0 | 96,203 (73.1) | 7,181 (82.6) |
| | 1 | 37629 (25.2) | 1553 (14.8) | | 1 | 35405 (26.9) | 1510 (17.4) |
| Age_55_60 | 0 | 115,723 (77.5) | 8,499 (81.0) | Age_55_60 | 1 | 31,055 (23.6) | 1,803 (20.7) |
| | 1 | 33667 (22.5) | 2000 (19.0) | | 0 | 100553 (76.4) | 6888 (79.3) |
| Age_60_65 | 0 | 117,429 (78.6) | 7,804 (74.3) | Age_60_65 | 0 | 102,445 (77.8) | 6,445 (74.2) |
| | 1 | 31961 (21.4) | 2695 (25.7) | | 1 | 29163 (22.2) | 2246 (25.8) |
| Age_65_70 | 0 | 128,816 (86.2) | 8,484 (80.8) | Age_65_70 | 0 | 114,162 (86.7) | 7,141 (82.2) |
| | 1 | 20574 (13.8) | 2015 (19.2) | | 1 | 17446 (13.3) | 1550 (17.8) |
| Age_70_75 | 1 | 14,688 (9.8) | 1,322 (12.6) | Age_70_75 | 0 | 120,403 (91.5) | 7,641 (87.9) |
| | 0 | 134702 (90.2) | 9177 (87.4) | | 1 | 11205 (8.5) | 1050 (12.1) |
| Age_75_80 | 0 | 138,519 (92.7) | 9,585 (91.3) | Age_75_80 | 0 | 124,274 (94.4) | 8,159 (93.9) |
| | 1 | 10871 (7.3) | 914 (8.7) | | 1 | 7334 (5.6) | 532 (6.1) |
| censor_1 | uncensored | 148,711 (99.5) | 10,462 (99.6) | censor_1 | uncensored | 130,784 (99.4) | 8,647 (99.5) |
| | censored | 679 (0.5) | 37 (0.4) | | censored | 824 (0.6) | 44 (0.5) |
| censor_2 | uncensored | 147,990 (99.1) | 10,414 (99.2) | censor_2 | uncensored | 129,944 (98.7) | 8,593 (98.9) |
| | censored | 1400 (0.9) | 85 (0.8) | | censored | 1664 (1.3) | 98 (1.1) |
| censor_3 | uncensored | 147,278 (98.6) | 10,375 (98.8) | censor_3 | uncensored | 129,138 (98.1) | 8,545 (98.3) |
| | censored | 2112 (1.4) | 124 (1.2) | | censored | 2470 (1.9) | 146 (1.7) |
| censor_4 | uncensored | 146,594 (98.1) | 10,340 (98.5) | censor_4 | uncensored | 128,290 (97.5) | 8,488 (97.7) |
| | censored | 2796 (1.9) | 159 (1.5) | | censored | 3318 (2.5) | 203 (2.3) |
| censor_5 | uncensored | 145,885 (97.7) | 10,292 (98.0) | censor_5 | uncensored | 127,443 (96.8) | 8,442 (97.1) |
| | censored | 3505 (2.3) | 207 (2.0) | | censored | 4165 (3.2) | 249 (2.9) |
| censor_6 | uncensored | 145,162 (97.2) | 10,245 (97.6) | censor_6 | uncensored | 126,651 (96.2) | 8,392 (96.6) |
| | censored | 4228 (2.8) | 254 (2.4) | | censored | 4957 (3.8) | 299 (3.4) |
| event_1 | 0 | 148,603 (99.5) | 10,407 (99.1) | event_1 | 0 | 130,411 (99.1) | 8,585 (98.8) |
| | 1 | 787 (0.5) | 92 (0.9) | | 1 | 1197 (0.9) | 106 (1.2) |
| event_2 | 0 | 147,857 (99.0) | 10,341 (98.5) | event_2 | 0 | 129,245 (98.2) | 8,488 (97.7) |
| | 1 | 1533 (1.0) | 158 (1.5) | | 1 | 2363 (1.8) | 203 (2.3) |
| event_3 | 0 | 146,999 (98.4) | 10,272 (97.8) | event_3 | 0 | 128,081 (97.3) | 8,408 (96.7) |
| | 1 | 2391 (1.6) | 227 (2.2) | | 1 | 3527 (2.7) | 283 (3.3) |
| event_4 | 0 | 146,081 (97.8) | 10,196 (97.1) | event_4 | 1 | 4,744 (3.6) | 394 (4.5) |
| | 1 | 3309 (2.2) | 303 (2.9) | | 0 | 126864 (96.4) | 8297 (95.5) |
| event_5 | 0 | 145,200 (97.2) | 10,125 (96.4) | event_5 | 1 | 5,881 (4.5) | 494 (5.7) |
| | 1 | 4190 (2.8) | 374 (3.6) | | 0 | 125727 (95.5) | 8197 (94.3) |
| event_6 | 0 | 144,330 (96.6) | 10,068 (95.9) | event_6 | 1 | 7,071 (5.4) | 579 (6.7) |
| | 1 | 5060 (3.4) | 431 (4.1) | | 0 | 124537 (94.6) | 8112 (93.3) |
| Covars | mean (sd) | 9.7 (7.3) | 13.4 (7.8) | Covars | mean (sd) | 6.9 (5.8) | 10.6 (6.1) |
| Total_covars: | 243 | | | Total_covars: | 203 | | |

**Supplementary Table 3** Concordance index for the Cox regression across the different data splits.

| Targets | Concordance Female | Concordance Female sd. | Concordance Male | Concordance Male sd. |
|---|---|---|---|---|
| | | Train | | |
| Dementia | 0.709 | 0.002 | 0.730 | 0.002 |
| Extrapyramidal | 0.747 | 0.004 | 0.751 | 0.004 |
| CVD | 0.672 | 0.001 | 0.635 | 0.001 |
| Renal | 0.808 | 0.002 | 0.780 | 0.002 |
| COPD | 0.684 | 0.001 | 0.622 | 0.002 |
| Liver | 0.663 | 0.003 | 0.672 | 0.002 |
| IBD | 0.612 | 0.002 | 0.595 | 0.003 |
| Cancer | 0.558 | 0.001 | 0.562 | 0.001 |
| Death | 0.799 | 0.001 | 0.772 | 0.001 |
| | | Valid | | |
| Dementia | 0.703 | 0.008 | 0.728 | 0.008 |
| Extrapyramidal | 0.739 | 0.014 | 0.755 | 0.013 |
| CVD | 0.670 | 0.004 | 0.636 | 0.003 |
| Renal | 0.798 | 0.008 | 0.770 | 0.007 |
| COPD | 0.679 | 0.005 | 0.626 | 0.006 |
| Liver | 0.664 | 0.011 | 0.682 | 0.009 |
| IBD | 0.617 | 0.009 | 0.603 | 0.011 |
| Cancer | 0.553 | 0.004 | 0.555 | 0.004 |
| Death | 0.799 | 0.003 | 0.768 | 0.003 |
| | | Test | | |
| Dementia | 0.702 | 0.004 | 0.734 | 0.004 |
| Extrapyramidal | 0.741 | 0.006 | 0.740 | 0.006 |
| CVD | 0.669 | 0.002 | 0.634 | 0.001 |
| Renal | 0.798 | 0.004 | 0.775 | 0.003 |
| COPD | 0.684 | 0.002 | 0.620 | 0.003 |
| Liver | 0.666 | 0.005 | 0.673 | 0.004 |
| IBD | 0.615 | 0.004 | 0.608 | 0.005 |
| Cancer | 0.552 | 0.002 | 0.560 | 0.002 |
| Death | 0.798 | 0.001 | 0.774 | 0.001 |

**Supplementary Table 4** Pearson correlation for Bayesian Cox estimates between females and males.

| | Targets | Pearson Correlation |
|---|---|---|
| 0 | Dementia | 0.657 |
| 1 | Ext.pyram. | 0.861 |
| 2 | CVD | 0.668 |
| 3 | Renal | 0.79 |
| 4 | COPD | 0.638 |
| 5 | Liver | 0.723 |
| 6 | IBD | 0.722 |
| 7 | Cancer | 0.459 |
| 8 | Death | 0.803 |

**Supplementary Table 5** Pearson correlation for ATE estimates between females and males.

| | Targets | Pearson Correlation |
|---|---|---|
| 0 | Dementia | 0.603 |
| 1 | Ext.pyram. | 0.205 |
| 2 | CVD | 0.629 |
| 3 | Renal | 0.46 |
| 4 | COPD | 0.577 |
| 5 | Liver | 0.22 |
| 6 | IBD | 0.22 |
| 7 | Cancer | 0.129 |
| 8 | Death | 0.785 |

### 2 Supplementary Figures

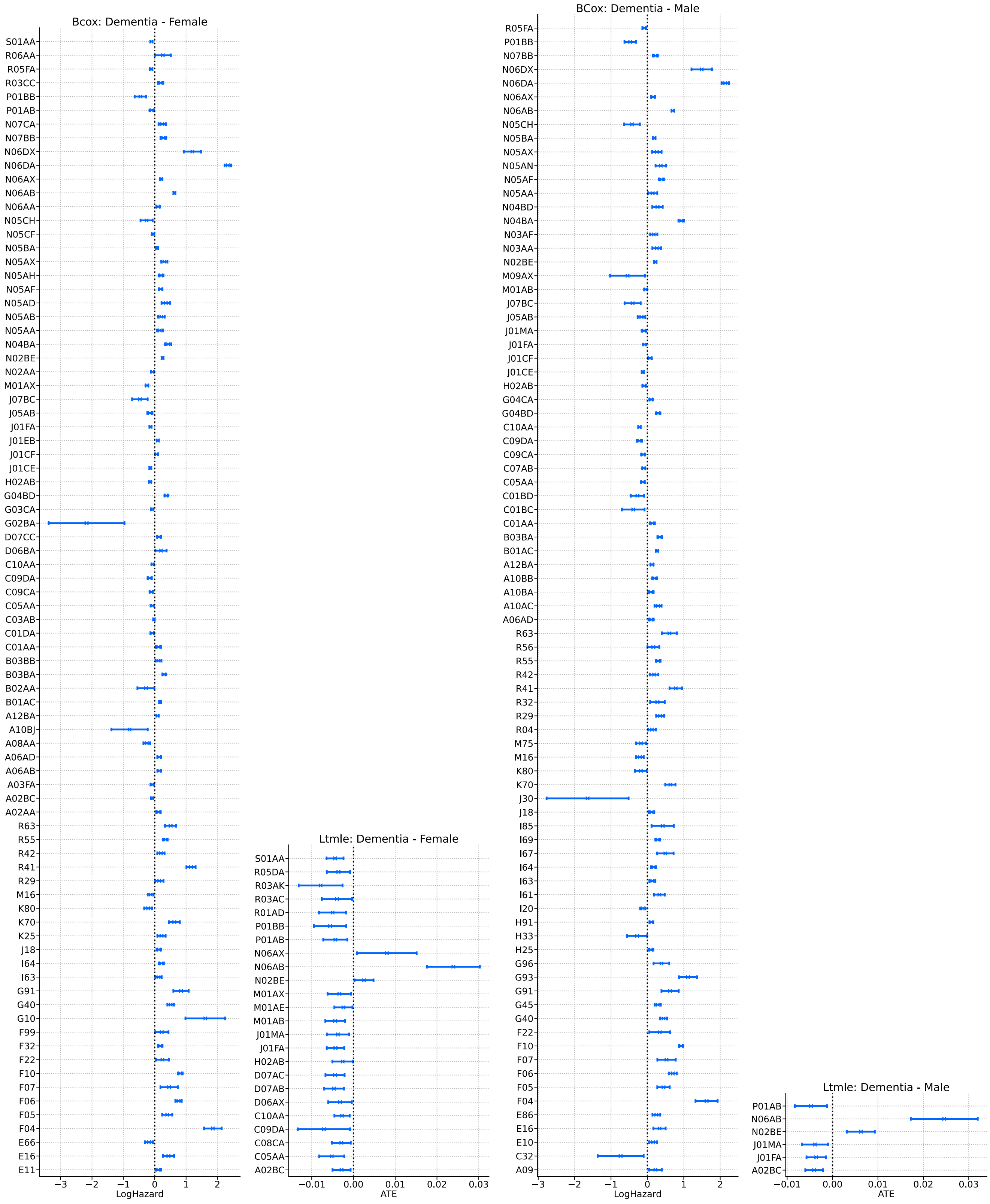


**Supplementary Figure 1** Significant 95% CI effect estimates for the Bayesian Cox regression and LTMLE for dementia.

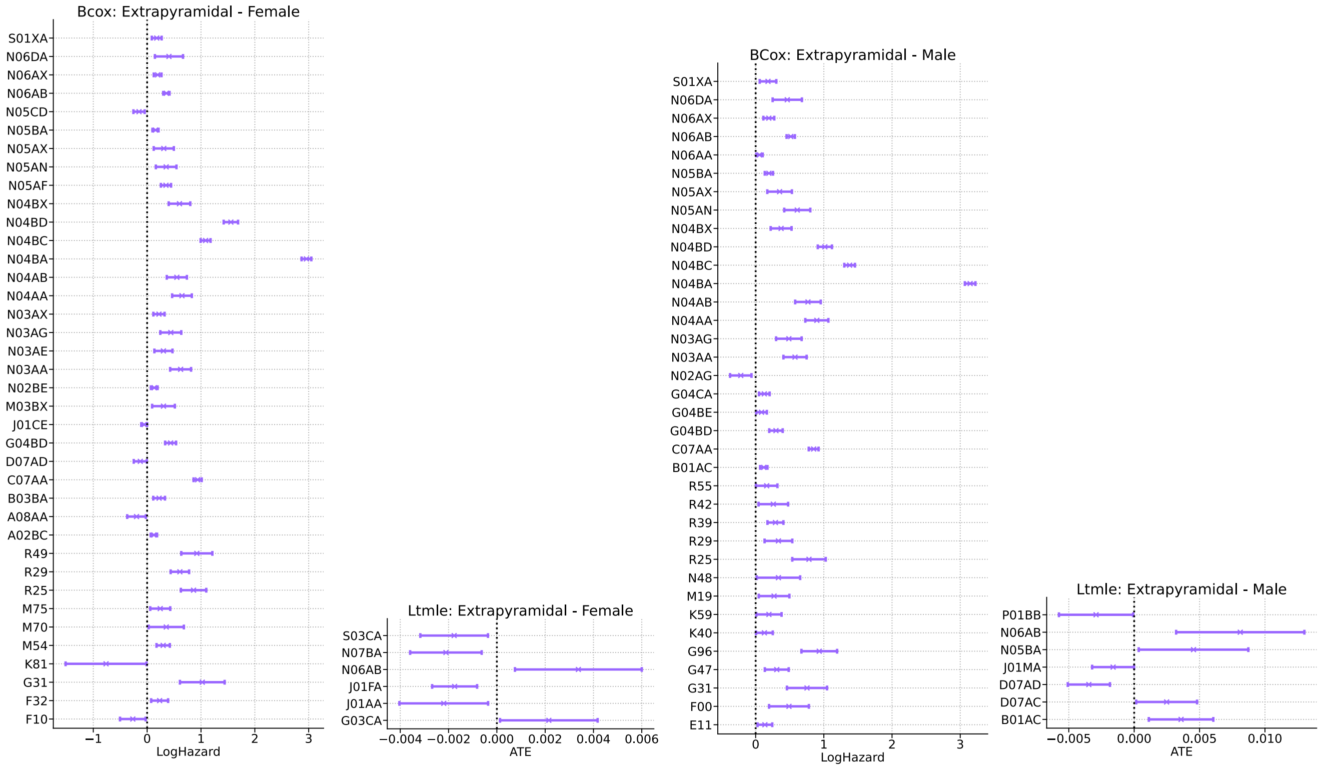


**Supplementary Figure 2** Significant 95% CI effect estimates for the Bayesian Cox regression and LTMLE for extrapyramidal disorders.

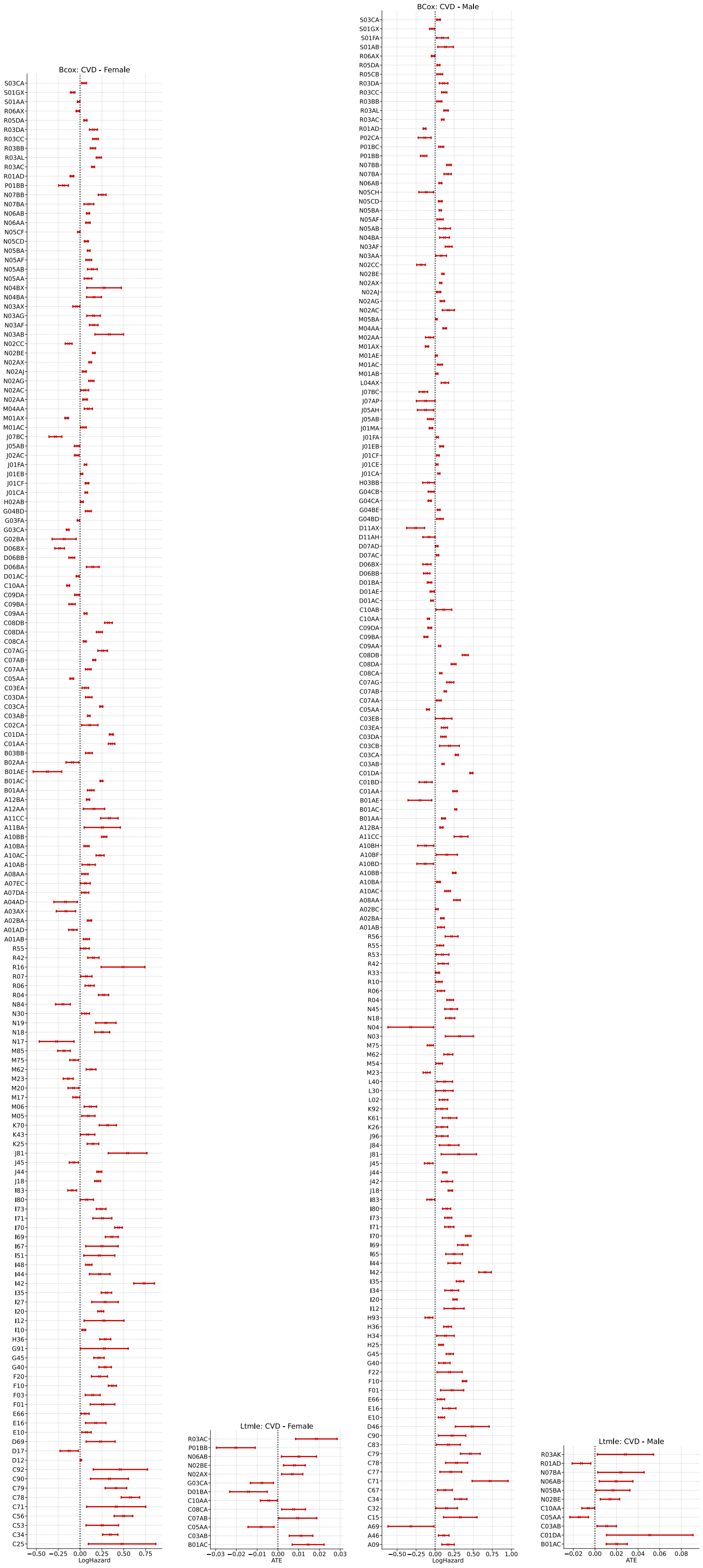


**Supplementary Figure 3** Significant 95% CI effect estimates for the Bayesian Cox regression and LTMLE for CVD.

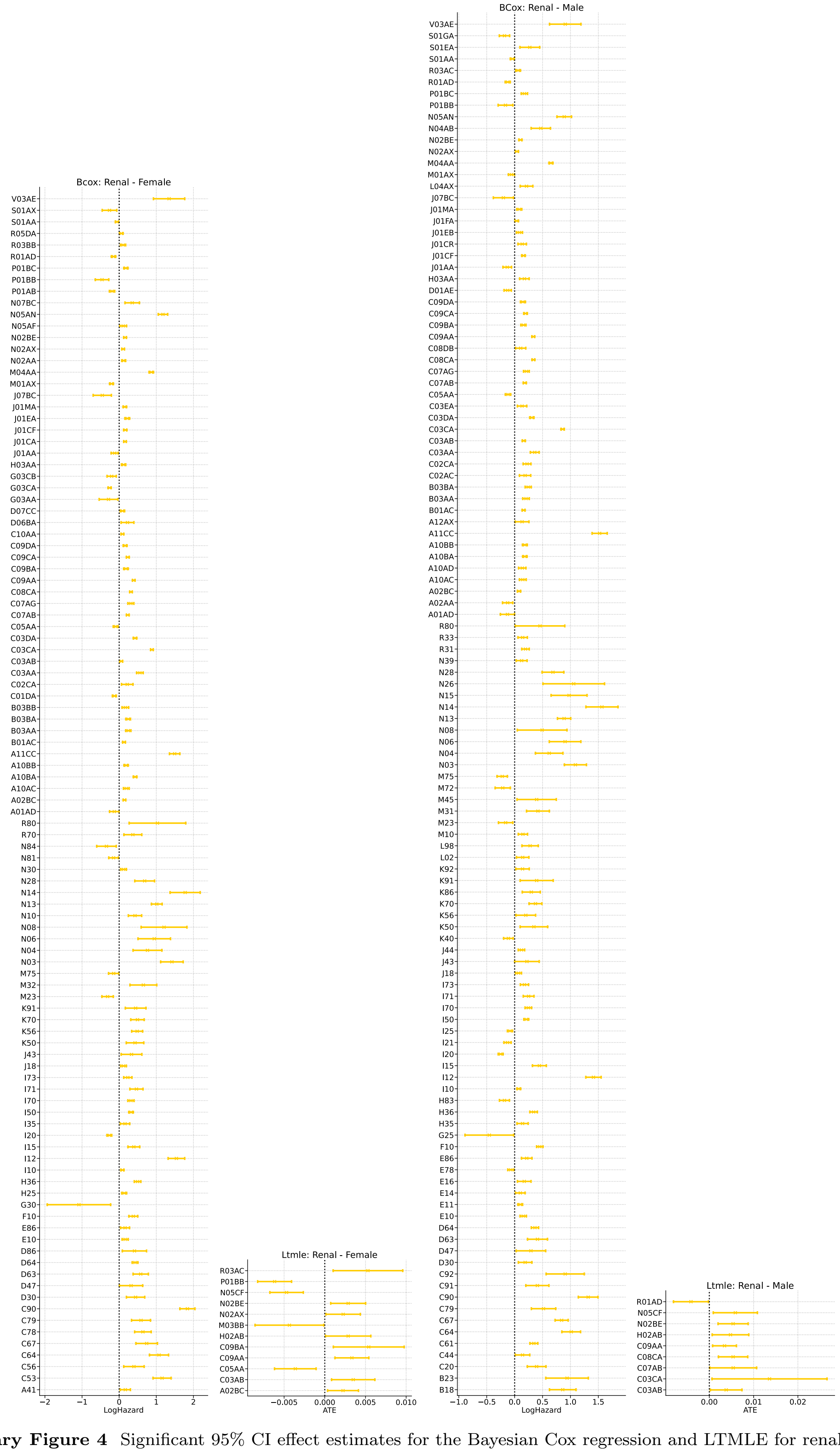

**Supplementary Figure 4** Significant 95% CI effect estimates for the Bayesian Cox regression and LTMLE for renal failure.

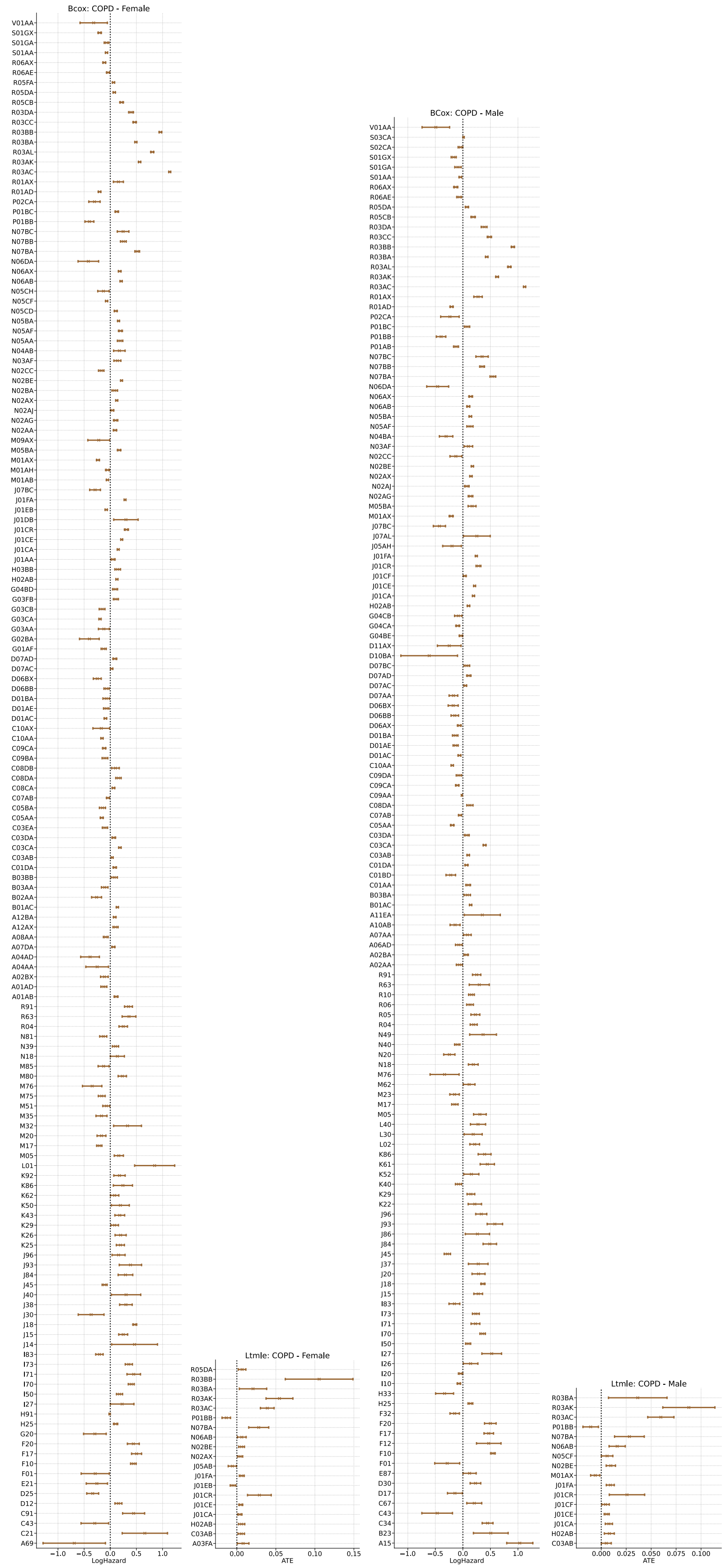


**Supplementary Figure 5** Significant 95% CI effect estimates for the Bayesian Cox regression and LTMLE for COPD.

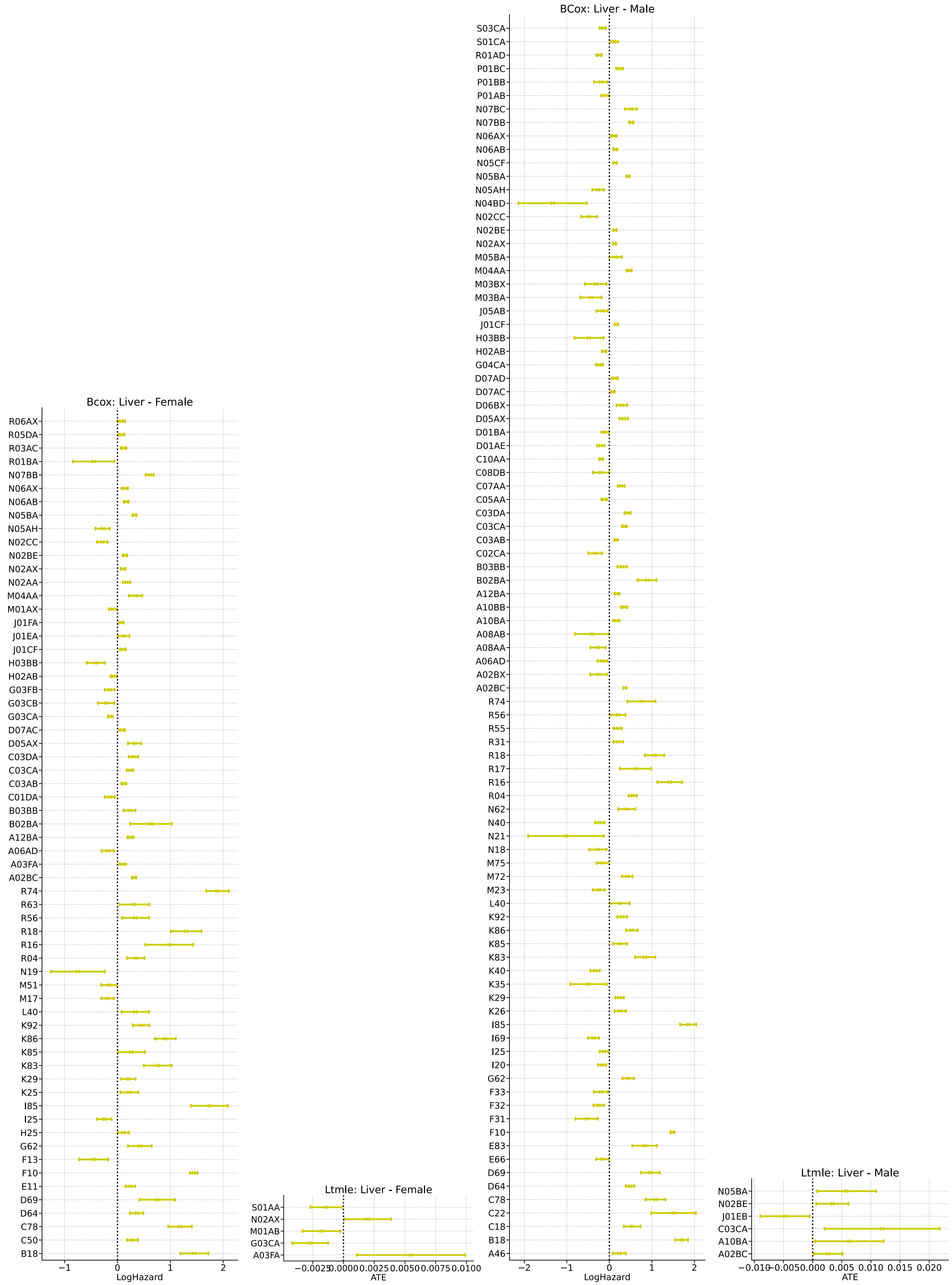


**Supplementary Figure 6** Significant 95% CI effect estimates for the Bayesian Cox regression and LTMLE for liver disease.

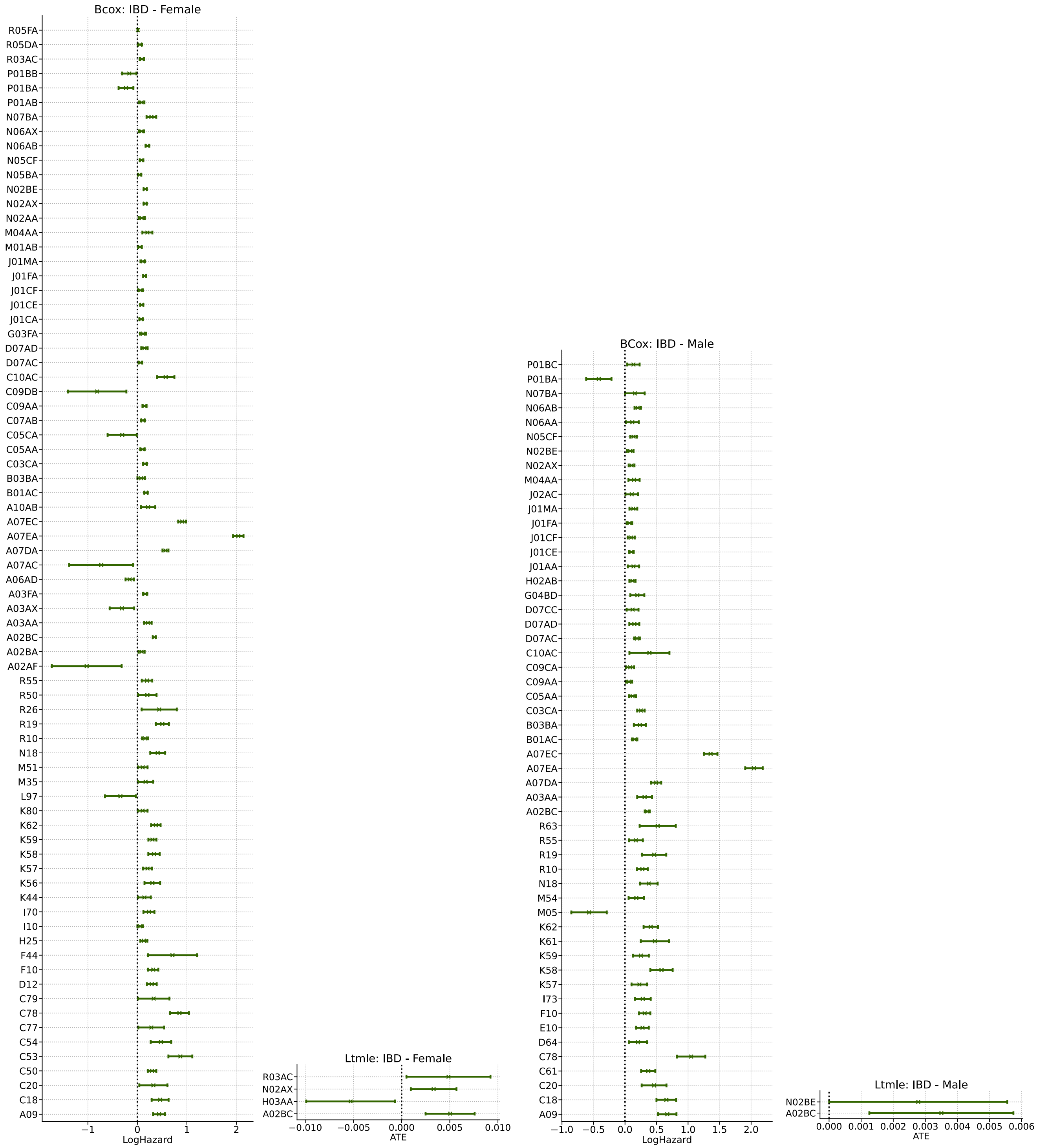


**Supplementary Figure 7** Significant 95% CI effect estimates for the Bayesian Cox regression and LTMLE for IBD.

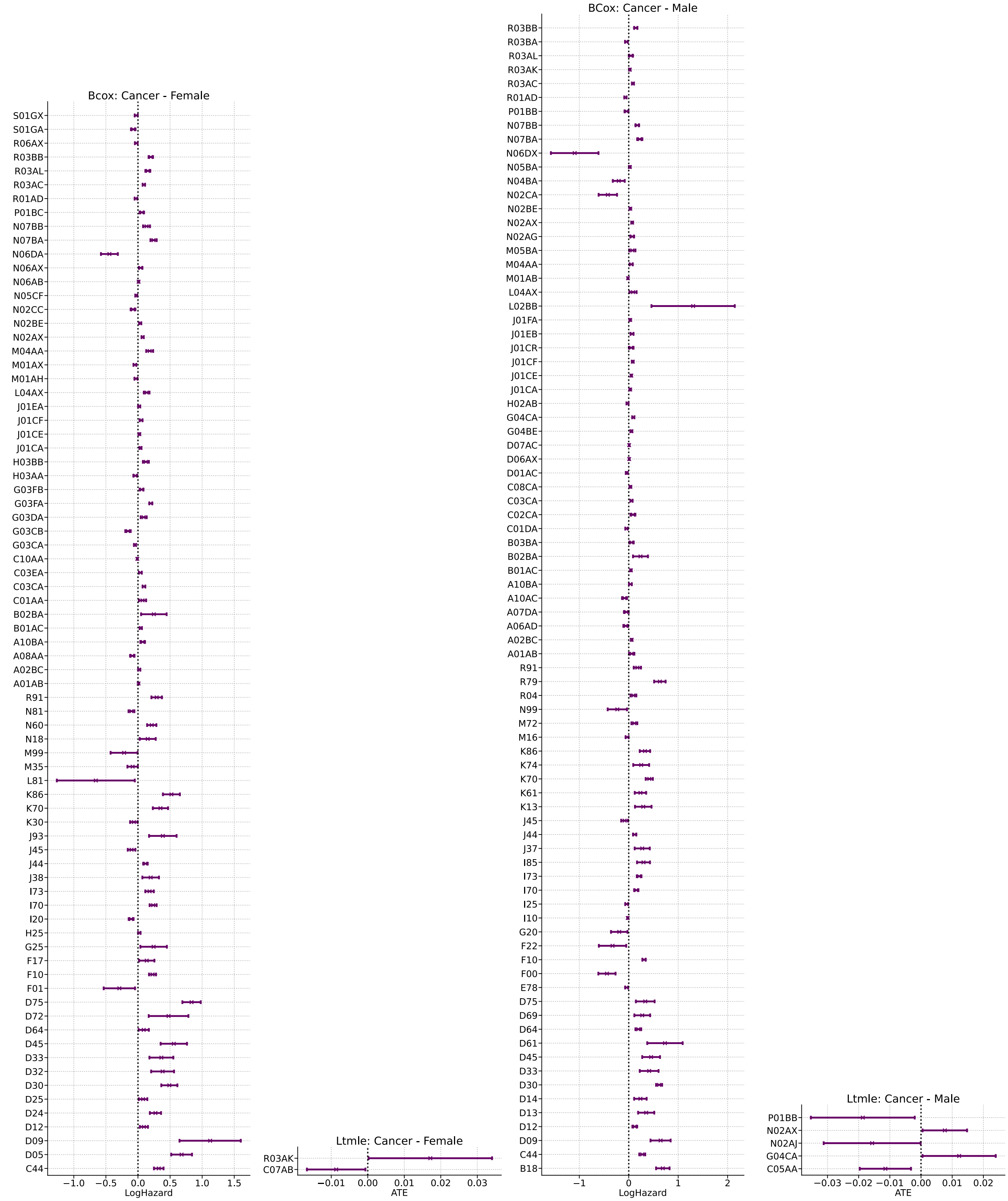


**Supplementary Figure 8** Significant 95% CI effect estimates for the Bayesian Cox regression and LTMLE for cancer.

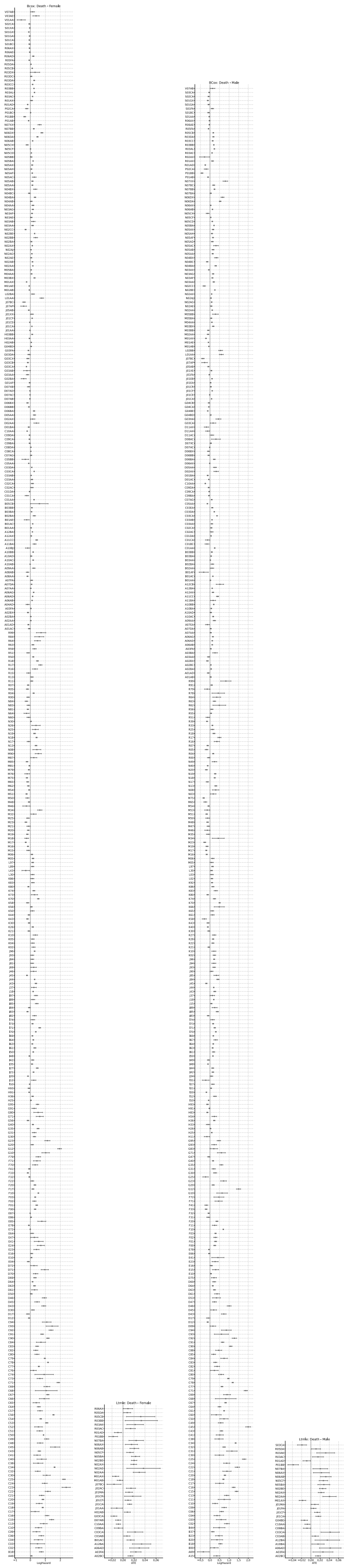

**Supplementary Figure 9** Significant 95% CI effect estimates for the Bayesian Cox regression and LTMLE for death.

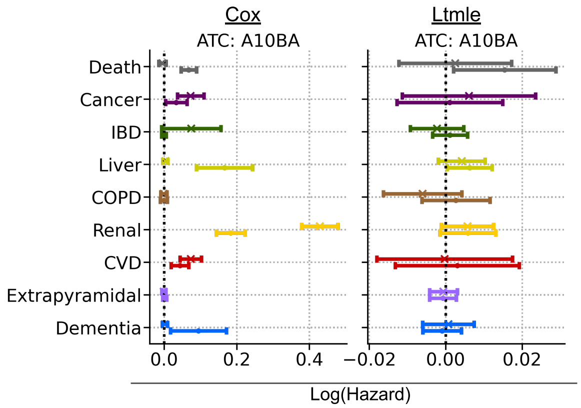


**Supplementary Figure 10** Effect estimates for ATC:A10AB from the observational Bayesian Cox regression on the left-hand side and the LTMLE analysis on the right-hand side for both sexes.

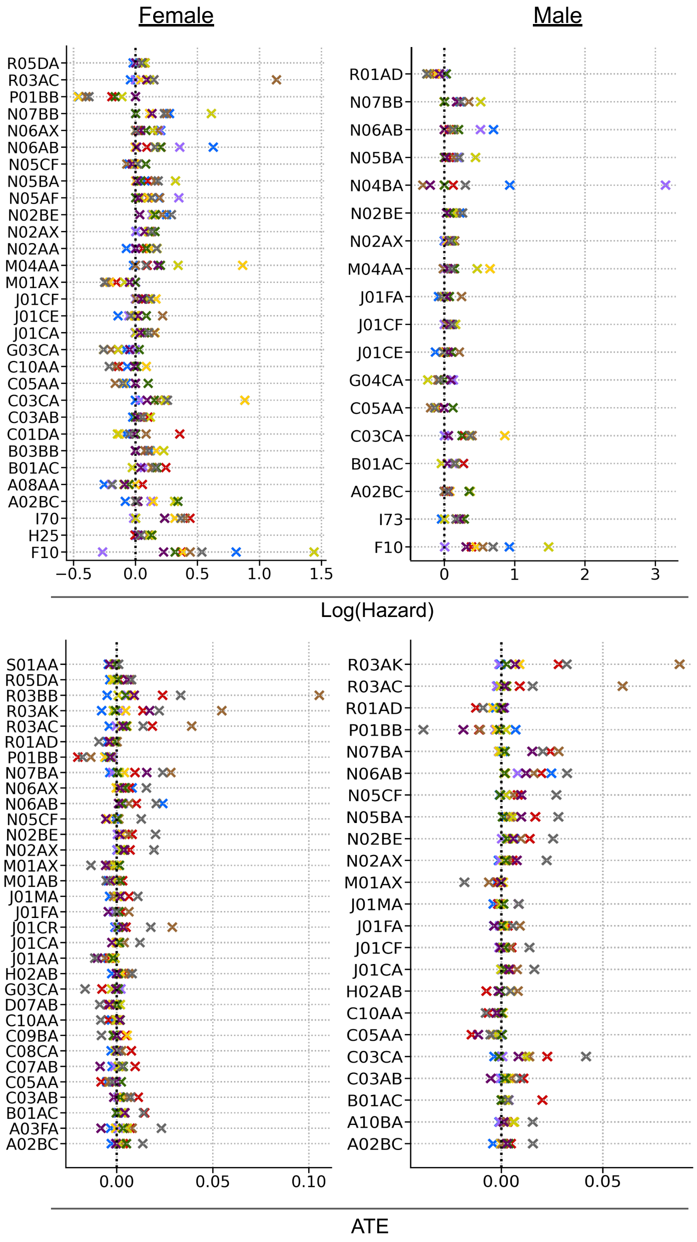


**Supplementary Figure 11** Effect estimates by sex for the observational Bayesian Cox regression and LTMLE analysis, summarized by ATC drug and showing all drugs with at least three significant effects.
